# Detection of antibodies to the SARS-CoV-2 spike glycoprotein in both serum and saliva enhances detection of infection

**DOI:** 10.1101/2020.06.16.20133025

**Authors:** Sian E. Faustini, Sian E. Jossi, Marisol Perez-Toledo, Adrian M. Shields, Joel D. Allen, Yasunori Watanabe, Maddy L. Newby, Alex Cook, Carrie R Willcox, Mahboob Salim, Margaret Goodall, Jennifer L. Heaney, Edith Marcial-Juarez, Gabriella L. Morley, Barbara Torlinska, David C. Wraith, Tonny V. Veenith, Stephen Harding, Stephen Jolles, Mark J. Ponsford, Tim Plant, Aarnoud Huissoon, Matthew K. O’Shea, Benjamin E. Willcox, Mark T. Drayson, Max Crispin, Adam F. Cunningham, Alex G. Richter

## Abstract

**Background:** Detecting antibody responses during and after SARS-CoV-2 infection is essential in determining the seroepidemiology of the virus and the potential role of antibody in disease. Scalable, sensitive and specific serological assays are essential to this process. The detection of antibody in hospitalized patients with severe disease has proven straightforward; detecting responses in subjects with mild disease and asymptomatic infections has proven less reliable. We hypothesized that the suboptimal sensitivity of antibody assays and the compartmentalization of the antibody response may contribute to this effect.

**Methods:** We systemically developed an ELISA assay, optimising different antigens and amplification steps, in serum and saliva from symptomatic and asymptomatic SARS-CoV-2-infected subjects.

**Results:** Using trimeric spike glycoprotein, rather than nucleocapsid enabled detection of responses in individuals with low antibody responses. IgG1 and IgG3 predominate to both antigens, but more anti-spike IgG1 than IgG3 was detectable. All antigens were effective for detecting responses in hospitalized patients. Anti-spike, but not nucleocapsid, IgG, IgA and IgM antibody responses were readily detectable in saliva from non-hospitalized symptomatic and asymptomatic individuals. Antibody responses in saliva and serum were largely independent of each other and symptom reporting.

**Conclusions:** Detecting antibody responses in both saliva and serum is optimal for determining virus exposure and understanding immune responses after SARS-CoV-2 infection.

**Funding:** This work was funded by the University of Birmingham, the National Institute for Health Research (UK), the NIH National Institute for Allergy and Infectious Diseases, the Bill and Melinda Gates Foundation and the University of Southampton.

## Introduction

COVID-19, caused by SARS-CoV-2, has resulted in millions of cases and more than 400,000 deaths around the world ^1^. Detection of active infection is routinely achieved by testing for viral RNA, but this approach cannot be used once symptoms have resolved. Antibody testing is useful to determine historic exposure to the virus, may provide insight into the immunological status of the individual and could be a measure of protection against re-infection.

The development of novel antibody tests requires a comprehensive understanding of the humoral response to a specific pathogen across the spectrum of disease caused by that pathogen. An important factor is the variable clinical presentation of infection that can influence the concentration of antibody induced within a subject. Understanding antibody responses in individuals with the lowest symptomatology will be of major importance for monitoring viral transmission within this SARS-CoV-2 pandemic ^2^. We have previously reported that asymptomatic seroconversion associates with lower levels of antibody to viral spike protein, which may complicate discriminating between asymptomatically infected individuals and those who were never infected^3^. Antigen choice and purity are other elements that can influence performance of the assay, not least by detecting cross-reactive antibodies induced by previous infection to other coronaviruses^4^. Therefore, the development of assays to detect low levels of anti-viral antibodies need to consider multiple variables in order to be of use in seroepidemiological studies.

Understanding the relationship between the varied clinical presentations of COVID-19 and the serological response that arises during and following infection will be of major significance in understanding the immunopathogenesis of disease and selecting appropriate treatments. This includes the degree of antigen recognition and the antibody subclasses involved. Little is known about the role of different antibody subclasses offering protection versus driving immunopathology in COVID-19. For instance, antibodies such as IgM, and the IgG subclasses IgG1 and IgG3 are efficient at activating complement, whereas IgA and IgG2 are not ^5^.

Most pathogens that enter via mucosal surfaces can induce immune responses within the mucosa and associated secondary lymphoid organs as well as systemic immunity in distant lymphoid organs, like the spleen. Systemic and mucosal immune responses can share significant overlap, yet the two immune systems are semi-autonomous. A clear example demonstrating the segregation of antibodies in the blood and mucosal compartments is the finding that in multiple myeloma patients, the monoclonal antibody and free light chains secreted by the malignant plasma cells is clearly detected in blood but not in saliva (unpublished observations). Nevertheless, other studies have shown that mucosal immunity can drive systemic responses demonstrating that an inter-relationship often occurs ^6, 7^. In the context of SARS-CoV-2 infection, the relationship between systemic and mucosal antibody responses is not completely understood. Assessing and understanding this aspect is important as it offers the opportunity to simplify testing through use of less invasive approaches, e.g. using saliva. Undermining the ready use of tests that examine salivary antibody levels, is that levels against specific pathogens can be a hundred to a thousand fold less than serum levels and thus fall under the level of detection of assays employed^8^. Mucosal antibody studies may also provide insights into the nature of post-infection protective immunity and help us understand the inter-relationship between systemic and mucosal immunity to the virus, which has applications for vaccine programs.

In this study, we report on the use of an antibody assay to detect antibodies in subjects with lower levels of SARS-CoV-2 specific-antibody. To do this, we examined responses to two well characterized proteins - the surface-exposed spike (S) protein that is a target of neutralizing antibodies and the nucleocapsid (N) protein, which is the most abundant viral protein. After identifying the optimal approach to maximize the signal:noise ratio, we then determined the relationship between antibodies in serum and saliva. This work will help accelerate the development of sensitive ELISA methods available to researchers and also inform on short and long-term immunity.

## Results

### Hospitalized patients induce robust responses to multiple SARS-CoV-2 antigens

To identify the antibody response to the virus, we tested sera against a range of viral antigens. There were three groups of subjects analyzed: Hospitalized subjects (HS, N=18), which included individuals that were admitted to the hospital and had confirmed SARS-CoV-2 infection by PCR; non-hospitalized convalescent (NHC, N=39) subjects, who were patients with confirmed SARS-CoV-2 infection by PCR but were not hospitalized; and asymptomatic non-hospitalized convalescent patients (AS, N=6), who were individuals without reported symptoms who gave a positive result for SARS-CoV-2 infection by PCR. As a negative control group, we used sera taken before 2019 (Pre-19, N=35). Further details can be found in supplementary table I. Initial studies focused on the two major targets of antibody responses: the viral S1 fragment and purified receptor binding domain (RBD) of spike (S) glycoprotein and nucleocapsid (N) proteins. As expected, strong IgG, IgA and IgM responses were detected to these proteins in all HS individuals with severe disease (Fig. 1). In contrast to the strong responses observed in severe cases, IgG, IgA and IgM responses were observed in the NHC subjects, and in some instances, responses were undetectable (Fig. 1B). At the first dilution there was some binding of IgG in Pre19 sera to N. Area Under the Curve (AUC) calculations confirmed that the highest response was observed in the group of HS for all the antigens tested (Fig. 1D). However, in AS and NHC, responses were reduced by half for IgG and at least to a fifth for IgA and IgM (Fig. 1D). Despite the lower antibody levels, IgG, IgA and IgM responses were detected in some asymptomatic individuals. Thus, simple non-optimized ELISAs readily detect antibodies to spike, RBD and N protein in sera from RT-PCR confirmed COVID-19 patients.

**Figure 1.**
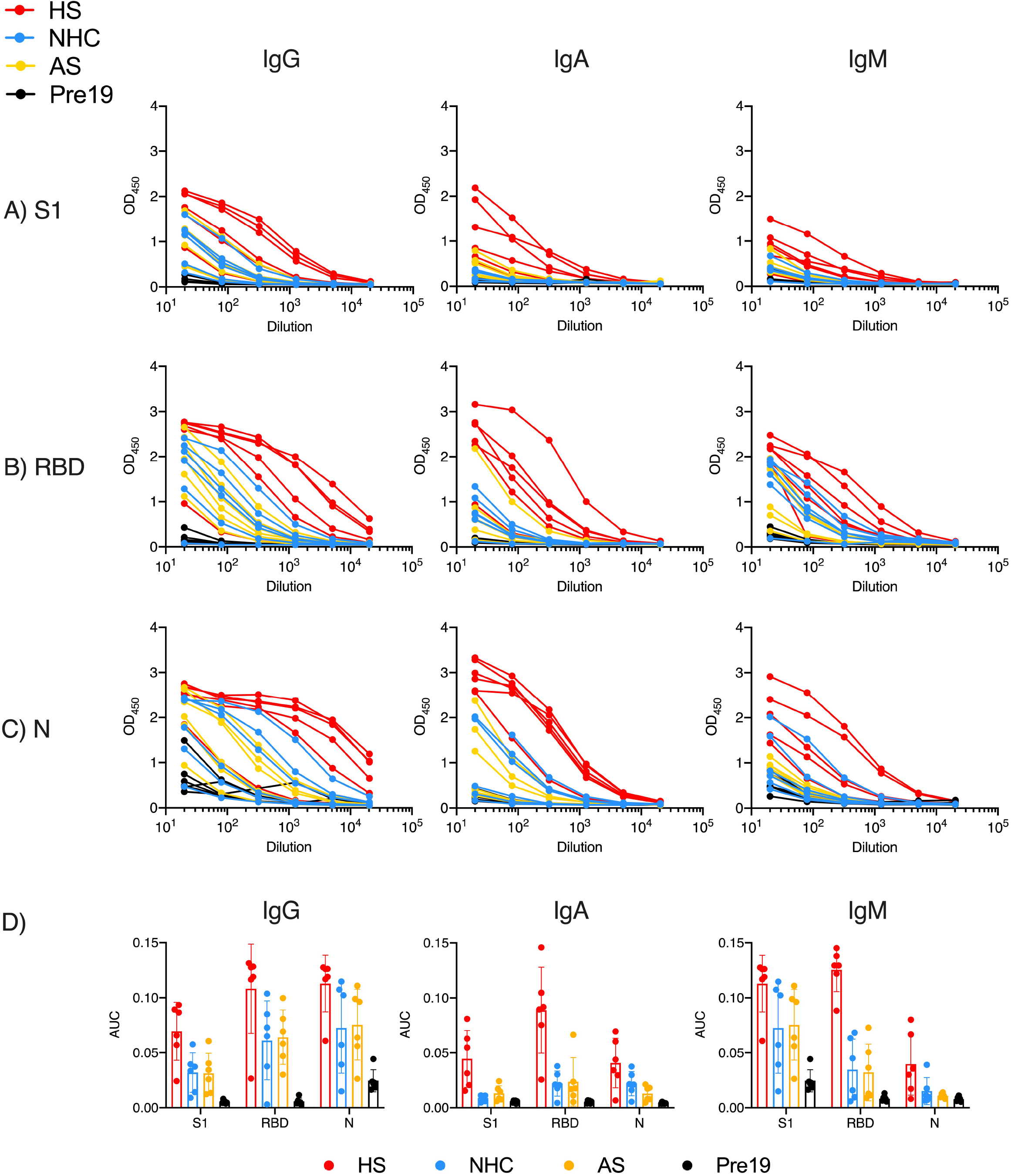
Hospitalised patients respond strongly to multiple viral proteins. Serological responses from hospitalised (HS, n=6), non-hospitalised convalescents (NHC, n=6), RT-PCR+ asymptomatic subjects (AS, n=6) or pre-2019 normal donors (Pre19, n=6) as determined by ELISA using HRP-labelled anti-IgG, IgA and IgM, against 0.1µg purified A) viral spike protein S1 fragment (S1), B) Receptor Binding Domain (RBD) or C) Nucleocapsid (N). D) Area Under the Curve (AUC) of responses shown in A-C. The mean ± standard deviation from the mean (SD) is plotted.

### Generation of soluble, native-like trimeric S glycoprotein

The use of RBD and S1 fragments within the assay was sufficient to detect antibodies in most individuals. Nevertheless, we hypothesized that these subunits may result in sub-optimal detection of antibodies in sera, particularly where titers were low, as these constructs both present intrinsically lower number of native epitopes and, in the case of the RBD, additionally display non-native epitopes hidden in the natively folded glycoprotein. Therefore, we produced soluble trimeric SARS-CoV-2 S glycoprotein. We expressed and purified recombinant SARS-CoV-2 S glycoprotein containing the previously described 2P-stabilized form, with a construct lacking the furin cleavage site, which minimizes S1/S2 subunit shedding, and is trapped in the pre-fusion conformation ^9^. The purity of the resultant SARS-CoV-2 S glycoprotein was confirmed by both SDS-PAGE (Figure 2A,B) and by mass spectrometry (reported in detail elsewhere ^10^). To ensure the SARS-CoV-2 was natively folded we performed surface plasmon resonance (SPR) between the purified S glycoprotein and its cognate receptor human angiotensin-converting enzyme (hACE2) (Figure 2C). We determined a K_D_ of 84 nM for hACE2 binding to SARS-CoV-2 S glycoprotein confirming the functionality of the purified glycoprotein.

**Figure 2.**
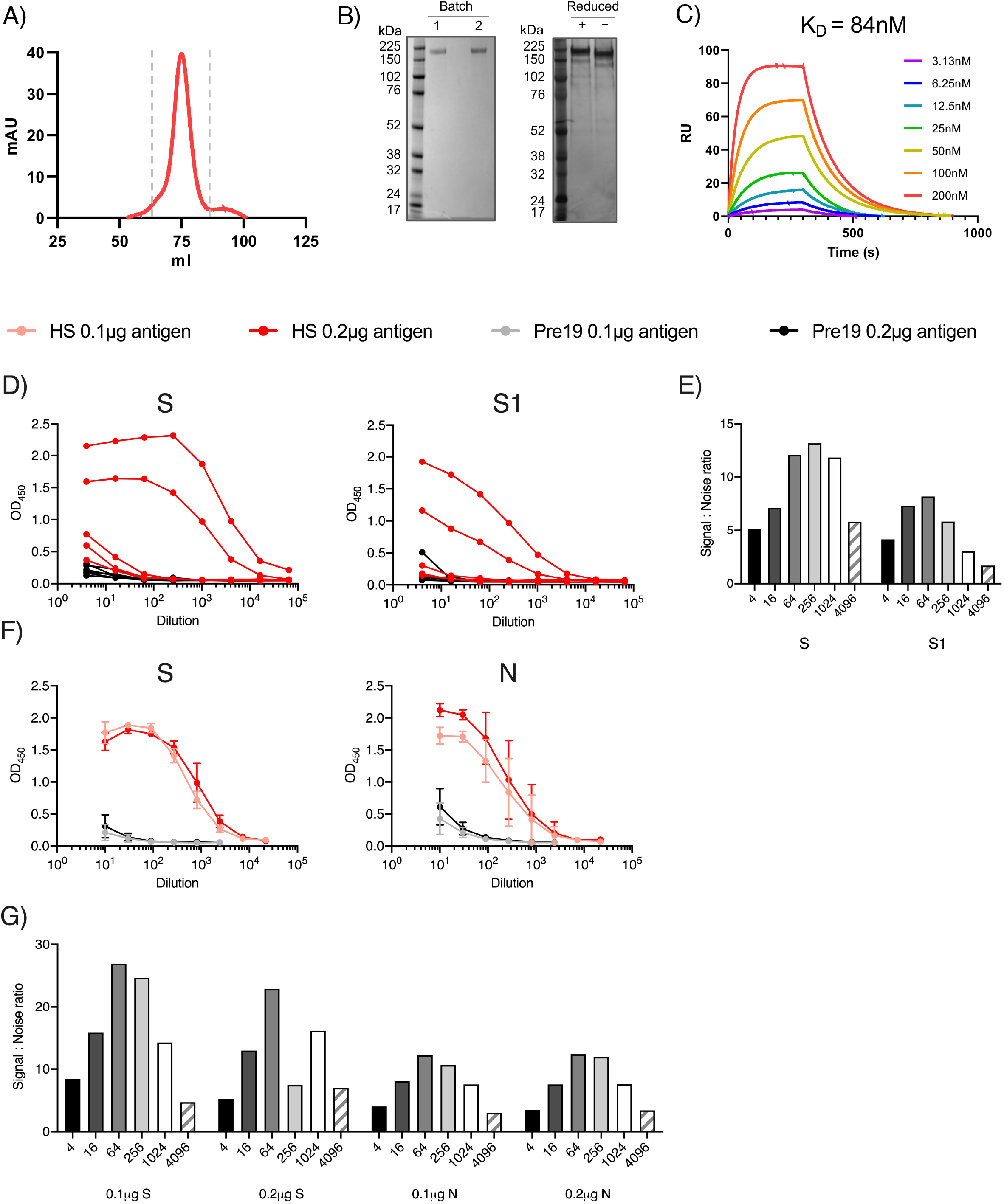
Stabilised, trimeric S antigen is a superior antigen to detect Ab in NHC. A) Size exclusion chromatogram (SEC) for SARS-CoV-2 S protein fractions collected for further use and denoted by dashed grey lines. B) Coomassie stained SDS-PAGE gel for two separate expressions of SARS-CoV-2 (left) and silver stain of batch one under reducing and non-reducing conditions (right). C) Surface plasmon resonance (SPR) characterizing the interaction between SARS-CoV-2 S protein and Ace2. The plotted lines represent the averages of three analytical repeats at each concentration. D) Serological responses from hospitalised (HS, n=9), or pre-2019 normal donors (Pre19, n=10) as determined by ELISA using HRP-labelled anti-IgG represented as absorbance values or E) Signal:noise ratio at each serum dilution against 0.1µg purified viral trimeric spike protein (S) or the S1 fragment (S1). F) Mean absorbance values of 6 sera per group against 0.1 or 0.2µg S or nucleocapsid (N). G) Signal:Noise ratio at each serum dilution against 0.1 or 0.2 µg of S or N. Error bars represent standard deviation from the mean (SD).

### Native-like, trimeric S antigen is superior to N to detect Ab in sera at higher dilutions

We then assessed whether changing multiple parameters within the assay could enhance the responses detected. By using the purified trimeric S glycoprotein we enhanced antibody detection compared to S1 protein, both in terms of the absolute OD_450_ values and in terms of the signal:noise ratio, and this was particularly notable as antibody became limiting (Fig. 2D,E). For example, signal:noise ratio when S glycoprotein was used was above 10 and only dropped when the sera was diluted to 1:4096. In contrast, signal:noise ratio for S1 remained lower than 10 at all dilutions tested. Increasing the amount of S glycoprotein per well from 0.1 to 0.2 µg/well, modestly enhanced the anti-S OD_450_ values (Fig. 2F), but overall provided no improvement of the signal:noise ratio (Fig. 2G). Similarly, higher concentrations of N did improve signal detection but had little difference to the signal:noise ratio, as background responses to control sera also increased (Fig. 2F,G). Thus, total trimeric S glycoprotein provides better discrimination to identify infected from non-infected individuals than S1 or N protein.

### Antibody subclass distribution does not differ depending upon the severity of disease

We then investigated the type of antibody response in patients by identifying the IgG subclasses generated against each antigen, and whether these responses varied to the different antigens. This matters beyond the seroepidemiological detection of infection because heavy chain use influences the effector function of antibodies. In HS and NHC, IgG1 and IgG3 were detected to S, RBD and N (Fig. 3A-C). However, AUC for IgG1 and IgG3 was lower in NHC compared to HS (Fig. 3D). IgG2 was largely undetectable regardless of the antigen or the origin of the sera. However, some samples were weakly positive for IgG4 to N but not S (Fig. 3C). The IgG1 response predominated against S or its RBD component. Thus, severe and less severe SARS-CoV-2 infections result in similar IgG antibody isotype switching profiles, although the extent of IgG1 and IgG3 isotype switching may differ between antigens.

**Figure 3.**
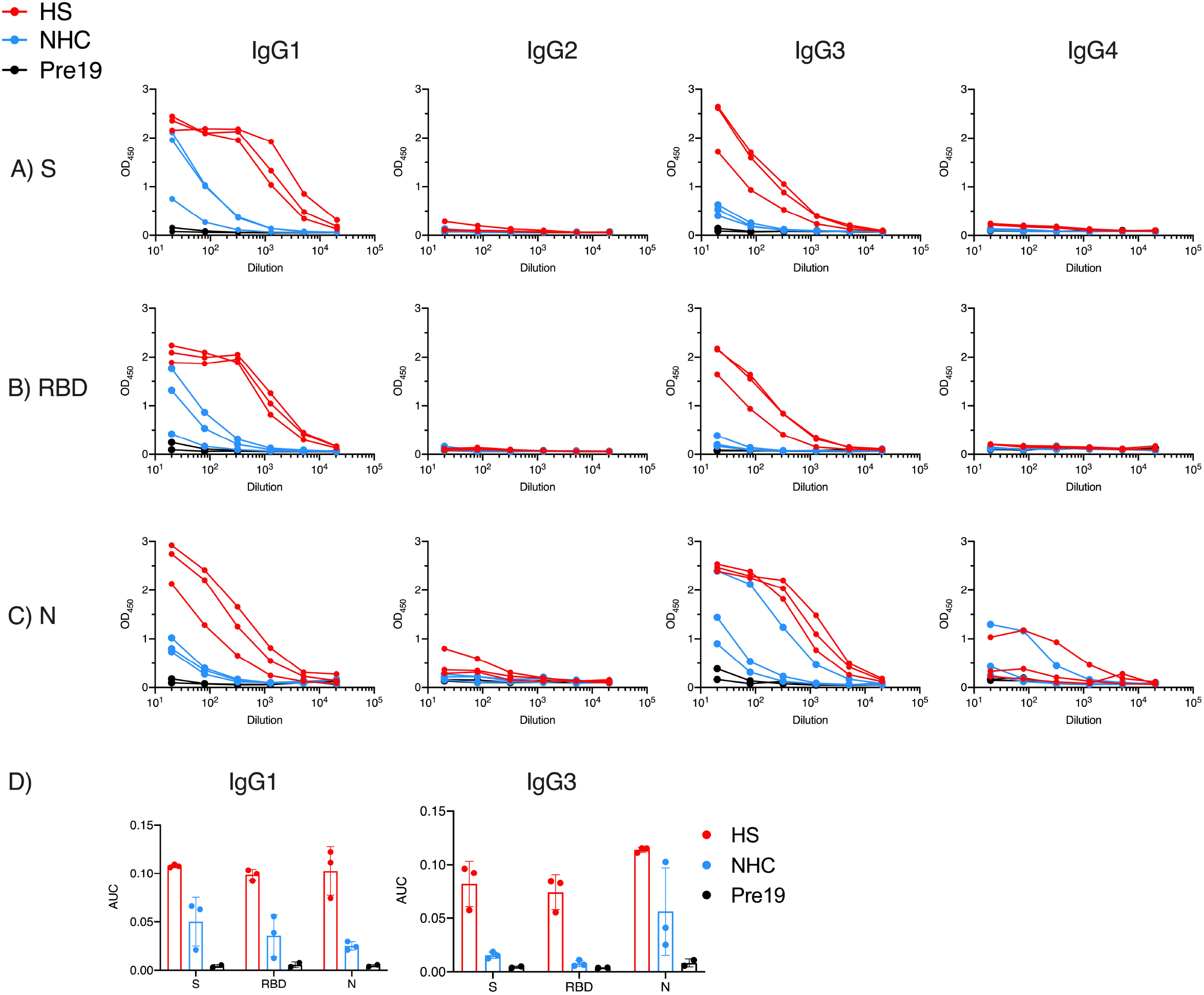
Antigen targeting and antibody isotypes do not differ depending upon the severity of disease. Serological responses from hospitalised (H, n=3), non-hospitalised convalescent (NHC, n=3) or pre-2019 donors (Pre19, n=3) as determined by ELISA using HRP-labelled anti-IgG1, IgG2, IgG3, or IgG4 against 0.1µg A) trimeric spike protein (S), B) Receptor Binding Domain (RBD) or C) Nucleocapsid (N). D) Area Under the Curve (AUC) of IgG1 and IgG3 responses as shown in A-C. The mean ± standard deviation from the mean (SD) is plotted.

### Combined detection of IgG, IgA and IgM enhances detection of antibody responses

The trimeric S glycoprotein was then used in the immunoassay to detect IgG, IgA and IgM in sera from NHC. All antibody isotypes were detectable in the same individuals (Fig. 4A). As a significant need in a test is to discriminate between infected individuals with low levels of antibody and non-infected individuals, we assessed whether combining anti-IgG, IgA and IgM (anti-GAM) secondary antibodies to detect all three isotypes could enhance signal detection. Merging secondary antibodies to detect anti-GAM responses provided a signal at each dilution that reflected the strongest signal from each of the three individual isotypes (Fig. 4A).

**Figure 4.**
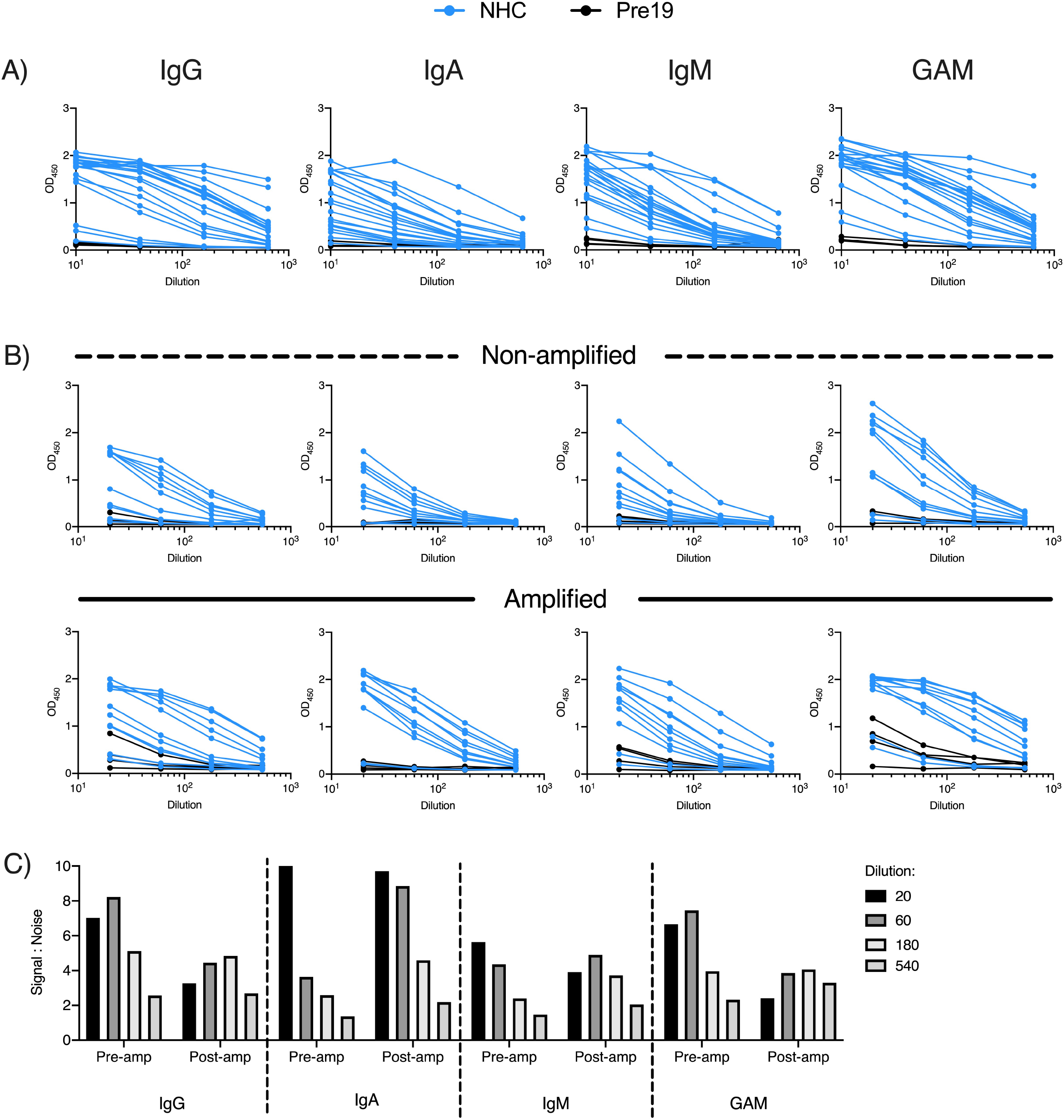
Combined detection of IgG, IgA and IgM enhances discrimination of infected and pre19 groups. Serological responses from non-hospitalised convalescents (NHC, n=20) or pre-2019 donors (Pre19, n=4) as determined by ELISA using HRP-labelled anti-IgG, IgA and IgM or combined GAM, against 0.1µg purified viral spike protein (S). A) Absorbance values of 20 NHC and Pre19 sera against S. B) Absorbance values of 14 low positive NHC sera with or without amplification, pre-2019 controls (n=4). C) Signal:noise ratio of multiple dilutions of anti-S IgG, IgA, IgM or GAM before and after amplification.

We examined if the IgG, IgA, IgM and GAM signals to S glycoprotein could be enhanced in a subset of sera from NHC subjects that had lower levels of antibody. To do this, we included an additional tertiary amplification step whereby after labelling primary antibodies with unconjugated mouse anti-human antibodies, HRP-conjugated anti-mouse immunoglobulins antibody was added. The inclusion of the amplification step enhanced the signal detected in nearly all samples, but had little effect on the signal:noise ratio for IgG (Fig. 4B,C). The enhanced signal detected for IgM and IgA (Fig 4B) resulted in higher signal:noise ratios for IgA and IgM, particularly at higher dilutions (S:N ratio at 1:540 dilution on pre-amplification 1.4 vs 2.2 post-amplification for IgA, and 1.5 vs 2.1 for IgM; Fig. 4C). Thus, this additional step is most beneficial to enhance signal detection when specific anti-S antibodies are present at lower concentrations.

### Anti-S, but not N, IgG, IgA and IgM responses are detectable in saliva from self-reported symptomatic subjects

Saliva is an easily accessible fluid that can be self-collected through a non-invasive procedure and could be beneficial for mass scale seroprevalence studies. Moreover, entry of the virus is via the upper respiratory tract and antibodies in saliva may provide a first barrier to entry at this point. Therefore, we investigated antibody responses in saliva from subjects who self-reported symptoms consistent with COVID-19 (SRSS). Initial experiments without amplification, assessing individual anti-IgG, IgA and IgM responses to S glycoprotein revealed the strongest signals were for IgA and IgG (Fig. 5A). In contrast, responses against N were not detectable above the Pre19 background responses (Fig 5B). As observed for sera, combining anti-IgG, IgA and IgM secondary antibodies typically bolstered the signal against S glycoprotein compared to when these secondary antibodies were used singly (S:N ratio at 1:32 dilution: IgG 1.53, IgA 1.40, IgM 1.1, Ig GAM 1.98; Fig. 5A). Nevertheless, the intensity of this signal was modest even when saliva was diluted just 1:8. Therefore, we assessed whether the amplification step used in Fig. 4 may improve the signal detected. Amplification of saliva antibodies increased the absolute OD_450_ values against S glycoprotein for IgG, IgM and GAM, but not for anti-N responses (Fig 5A, B). Amplification improved the signal:noise ratio most for anti-S glycoprotein IgG and IgM antibodies, with little or no enhancement of the ratio for IgA (Fig. 5C), particularly when the saliva was diluted. Adding the amplification step resulted in better signal:noise ratios for GAM at higher concentrations (S:N ratio at 1:32 for GAM was 1.98 pre-amplification vs 2.4 post-amplification; Fig. 5C). Thus, anti-S glycoprotein antibodies can be consistently detected in saliva from SRSS individuals.

**Figure 5.**
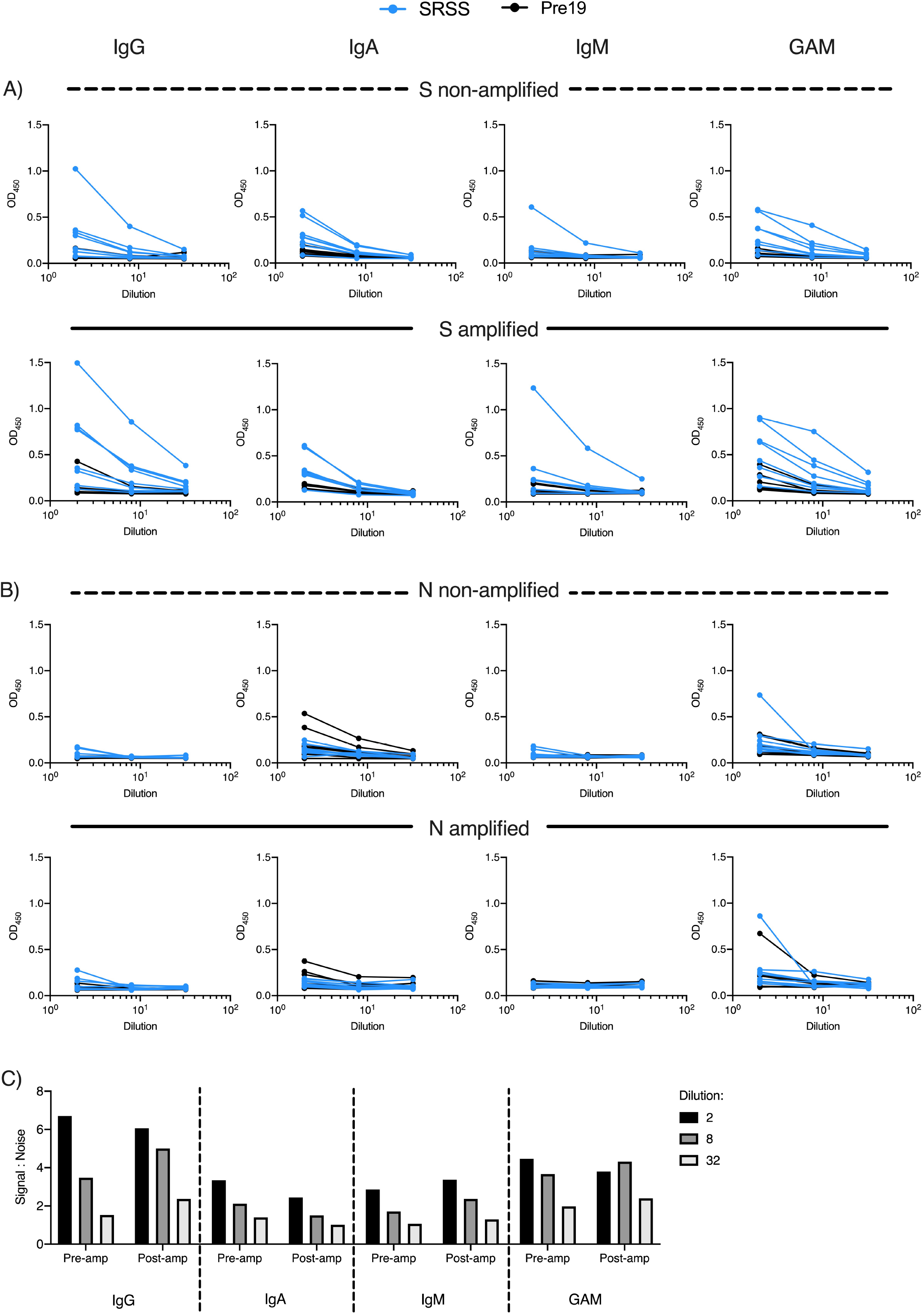
Amplification of antibody responses in saliva enables detection of S-specific, but not N-specific IgG, IgA and IgM. Salivary antibody responses from self-reported symptomatic subjects (SRSS, n=8) or pre-2019 negative controls (Pre19, n=4) as determined by ELISA using HRP-labelled anti-IgG, IgA and IgM or combined GAM, against 0.1µg purified viral spike protein (S). A) Absorbance values of NHC and Pre19 saliva against S before amplification and B) after amplification. C) Absorbance values of the same saliva samples against N before or D) after amplification. E) Signal:noise ratio of multiple dilutions of anti-S salivary IgG, IgA, IgM or GAM before and after amplification.

### Antibodies to S glycoprotein in saliva can be detected independently of serum responses

We have little understanding of the immune response that develops to SARS-CoV2 in the mucosa and its relationship to serum. Therefore, the relationship between antibody responses was assessed in 39 matched saliva (1:2 dilution) and serum (1:40 dilution) from a cohort of health-care workers that were recruited from the University Hospitals Birmingham NHS Foundation Trust 8. All of these subjects were asymptomatic and PCR-negative at the time of sampling. Of these 39 subjects, 18 reported never having COVID-19-associated symptoms, and thus were described as asymptomatic and 21 had noted symptoms at some stage in the past that were consistent with COVID-19. Within this population there are likely to be a mix of individuals who have never been infected, infected and asymptomatic and infected and recovered.

We plotted the OD450 of the 1:2 dilution of saliva for the amplified anti-GAM to S glycoprotein against the OD450 of the 1:40 dilution for sera. In the whole group, there was a modest correlation between the OD450 of saliva and sera (R = 0.389, Fig. 6A). Nevertheless, the individuals with sera giving the highest OD450 also had the highest saliva OD450. Splitting the group by whether they previously recorded symptoms or not, did not influence the distribution of responses in saliva or serum (Fig. 6B). The results were also categorized into whether individuals gave a negative or positive result based on being lower or higher respectively of a cut-off (mean + 3 SD of 6 saliva and 8 sera from pre19 samples). There were 13 (33.3%) subjects with no response to S glycoprotein and 9 (23.1%) positive in both saliva and serum. Six (15.4%) subjects were found with positive saliva but not serum and 11 (28.2%) with positive serum but not saliva. The agreement between the results was assessed with kappa statistics. Binary agreement between the OD450 for serum and saliva was poor (κ = 0.13 ±0.15 SE). Therefore, assessment of both serum and saliva increases the detection of individuals who have antibody responses to SARS-CoV-2,

**Figure 6.**
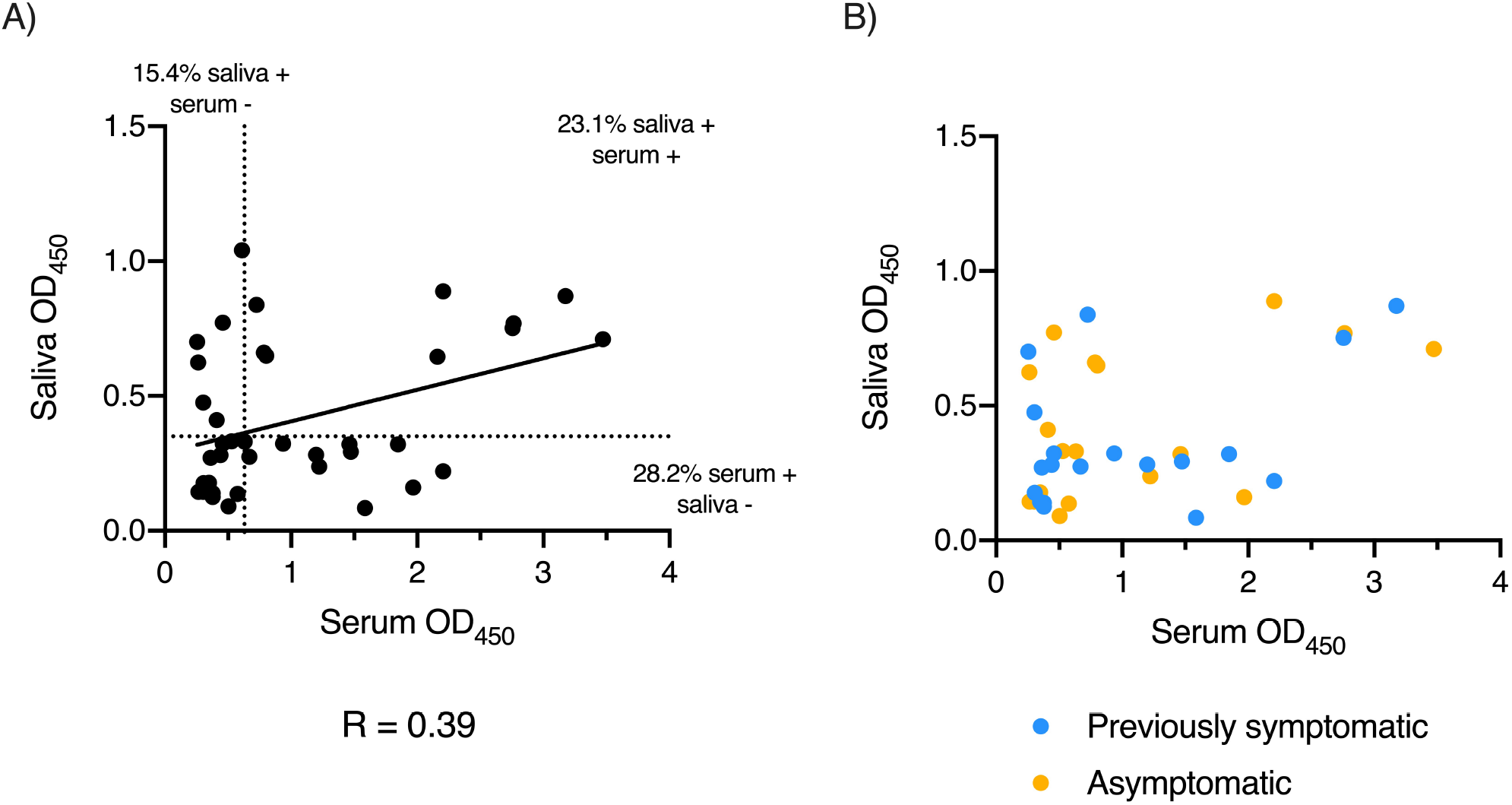
Serum and salivary anti-SARS-CoV-2 antibody responses do not always correlate. Absorbance values of paired serum diluted 1:40 and saliva diluted 1:2 from RT-PCR negative health care workers (n= 39) that were asymptomatic at the time of testing serum, as determined by ELISA using combined HRP-labelled anti-IgG, IgA and IgM (GAM) against 0.1µg trimeric spike protein (S) with signal amplification. A) Correlation between paired serum and saliva absorbance values with percentages of samples positive for anti-S antibodies in either serum, saliva or both. Positivity was determined by cut-offs for each fluid (dotted lines) based on the mean + 3 standard deviations of 6 pre-2019 (Pre19) negative samples. Solid line represents simple linear regression of all samples. B) Correlation of the same paired serum and saliva absorbance values coded by self-reported historic SAR-CoV-2-like symptoms (blue) or no historic symptoms (yellow).

## Discussion

The recent pandemic caused by SARS-CoV-2 has caused significant morbidity and mortality around the world. The need to be able to identify those who have previously had a SARS-CoV-2 infection has resulted in the development of immunoassays that are designed to measure antibodies as a signature of exposure, and there is a need to make these as sensitive and specific as possible. Antibody assays have also shown potential in diagnosing SARS-CoV-2-associated complications, such as helping diagnose children who present with PIMS-TS (Pediatric Multisystemic Inflammatory Syndrome), yet are PCR-negative for virus, and to define seroprevalence in symptomatic and asymptomatic health care workers in a hospital setting^3, 11, 12^. These real-world examples of the use of this assay emphasize the potential for these studies to aid in diagnosis and immunosurveillance. If post-infection complications arise from COVID-19, then the availability of high-quality assays to detect prior infection will be of obvious benefit. Collectively, these points add to the wealth of evidence supporting the benefit and value of antibody assays in the current crisis.

Our method focused on implementing different approaches to find the best signal:noise ratio. To do this, we used sera obtained from subjects prior to 2019, and sera from patients with confirmed RT-PCR infection with different severities of disease. Within these sera from infected patients, we were particularly interested in enhancing the signal:noise ratio in sera that either contained lower levels of antibody or the ratio when sera were diluted. RBD, S1 and N were excellent at detecting antibodies in sera from subjects with severe COVID-19, but were not as good as purified, whole, trimeric S when antibody levels were more limiting. This is unlikely to be due to the source of antigen preparations as we compared commercially purchased N, as well as antigens made within our own facility. Wider use of S glycoprotein in assays has been facilitated by recent improvements that have increased yields as much as 10-fold ^13^. Due to higher protein yields obtained after purification from culture, some studies suggest the use of RBD as a first screening test ^14^. This is likely to be useful for detecting responses in those with higher levels of antibodies. However, RBD and S1 have a more limited set of native epitopes present than whole native S glycoprotein and, at least in our studies, this potentially negatively affected the signal:noise ratio. It could be hypothesized that a greater diversity of epitopes is beneficial as there is more opportunity to capture a wider range of antibody specificities, and this may be more important when antibodies are rarer in a patient sample. Using the trimeric S glycoprotein may also help detect antibodies that block both binding and viral entry, which may offer additional insights than examining responses to RBD alone. Coupled with this is the presence of non-native epitopes in the RBD that are not present in the whole native trimeric S glycoprotein, and the purity of the S glycoprotein generated, with the glycan profiles offering an additional quality assurance step in determining purity. Moreover, the use of mammalian expression systems reduces the likelihood of detecting antibodies to antigens that have been encountered during normal exposure to pathogens, which can occur when organisms such as *E. coli* are used in the generation of recombinant N production. In our studies, we consistently found higher background responses in sera obtained prior to 2019 to N, the antigen used in the two chemiluminescence methods approved in the U.K. for detecting anti-SARS-CoV-2 antibodies^15^.

In our modified ELISA method, S protein gave a better discrimination between SARS-CoV-2 positive and negative samples than N. One concern of using the extracellular region of S in assays has been the risk of detecting antibodies that are cross-reactive between SARS-CoV-2 and SARS-CoV ^16^. Nevertheless, there are only around 8000 known SARS-CoV infected patients from 2003 and there are now well over 7 million individuals infected with SARS-CoV-2, so the proportional chance of detecting one of these patients is low. Furthermore, there is a similar level of identity of N between these viruses and thus, these are factors that can possibly influence responses to both SARS-CoV viruses. The level of amino acid conservation of S with other human pathogen members of the coronavirus family, such as HCoV-OC43 and HCoV-229E, is lower than to SARS-CoV. These two strains are thought to account for 5-30% of upper respiratory tract infections and most individuals who are infected with these strains do so from an early age ^17, 18^. Additionally, the low identity of N and S from SARS-CoV-2 with these viruses means cross-reactivity of antibodies to these proteins is minimal ^4^.

Studies in cohorts of SARS-CoV-2 patients show that antibody responses develop at different rates depending on the severity of the disease. In general, antibody titers are higher in patients with critical or severe disease when compared to those with milder disease ^19, 20^. Likewise, in our investigations, we found that samples from hospitalized patients had stronger IgG, IgA and IgM responses against S1, RBD and N antigens. The important finding from these responses is that it is relatively simple to detect antibodies in patients with severe disease and that focusing on detecting responses in those who have much less severe disease or are asymptomatic may be advantageous for maximizing the sensitivity of an antibody test. When the IgG subclasses were evaluated, IgG1 and IgG3 were the most abundant in all samples tested, and they were also higher in hospitalized patients than in those with mild disease. This is important because it has been suggested that antibodies may play a role in pathogenesis, including the possible role of IgG1 as a mediator of acute lung injury in COVID-19 ^21^. Of interest, the IgG1 signal to S was consistently stronger than that of IgG3, whereas to N a predominance of IgG1 or IgG3 was less clear. It would be a valuable study to examine IgG responses longitudinally in patients with different presentations of SARS-CoV-2 and examine how this relates to different presentations of infection.

Self-collected saliva is an attractive way to evaluate antibody prevalence due to the accessibility of the sample and non-invasiveness of the procedure, but fewer studies have explored this as a route to detect infected individuals. One reason for this is that it can be more challenging to detect antibodies in saliva. The use of an additional tertiary antibody incubation with an HRP-conjugated goat anti-mouse Ig to amplify the signal of the bound IgG, IgA or IgM resulted in an enhanced detection of anti-S, but not anti-N responses in individuals that had weak saliva antibody responses. The lack of detection of anti-N Ig GAM responses in saliva was unexpected, as it was readily detectable in serum and has been reported in saliva from more severe COVID-19 cases ^22^. The reasons for our lack of detection are unclear but may reflect greater partitioning of antibody responses between mucosal and systemic sites than appreciated previously, or the lack of local exposure in the oral cavity to N compared to other sites, or other unknown reasons.

When we assessed matched serum and saliva samples from a cohort of health care workers that was part of a study recently published ^3^, there was a minimal correlation between anti-S antibodies in serum and saliva. Nevertheless, many individuals were only single positive in serum or saliva. Thus, relying on one of these fluids to determine exposure may significantly underestimate true levels of exposure. Moreover, this discrepancy may have implications for our understanding of what forms a protective antibody response, and whether antibodies in one or more sites are required for optimal protection. In other studies from our group, this compartmentalization of antibody responses between serum and saliva has been observed in the context of anti-pneumococcal vaccine responses^23^, and has been seen in patients with multiple myeloma, who can have high serum levels of paraprotein which is absent in saliva (Heaney, J. L. J. unpublished data). This difference between mucosal and systemic antibody responses may be due to antibodies secreted by local plasma cells in the buccal cavity ^8, 24^. Therefore, the kinetics of antibody induction in distinct mucosal sites still needs further examination

Therefore, standard ELISA methods based on high-quality S protein can be modified to readily detect antibody responses in serum and saliva from severe, mild and asymptomatic COVID-19 infections. This method may serve as an important tool for assessing both short and long-term humoral immunity for community-acquired COVID-19 infections and understanding the nature of natural and vaccine-induced protective responses to SARS-CoV-2 infection

## Methods

### Patient cohorts and ethical review

Paired serum and saliva samples were collected from health care workers at University Hospitals Birmingham NHS Foundation Trust as part of the CoCo study. The study was approved by the London - Camden & Kings Cross Research Ethics Committee reference 20/HRA/1817. Pre2019 negative controls were recruited as part of a University of Birmingham study – reference ERN_16-178. All participants in both studies provided written, informed consent prior to their enrolment. Surplus serum samples from individuals with a history of PCR proven SARS-CoV-2 infection at University Hospitals Birmingham NHS Foundation Trust and the Immunodeficiency Centre for Wales were fully anonymized and used for assay development and quality assurance.

### Sample Collection

Serum samples were obtained from whole blood after centrifugation at 3500 RPM for 5 minutes and were stored at −20 °C until used in the assay. Whole saliva samples were collected by passive dribble into 50 ml saliva collection tubes for a timed period of 4 minutes. Samples were centrifuged (4000 RPM for 10 minutes) to separate cells and insoluble matter and the supernatant was removed and stored at –20°C until use. On the day of assay samples were thawed and microcentrifuged (10000 RPM for 10 minutes).

### Antigens used and SARS-CoV-2 spike protein production

The S1 subunit of the S glycoprotein and Nucleocapsid (N) proteins were obtained from the Native Antigen Company (UK). The Receptor Binding Domain (RBD) was prepared at the University of Birmingham. Briefly, the sequence encoding RBD (amino acids 319-541) of the SARS-CoV2 spike protein including a C-terminal hexahistidine tag in the pCAGGS mammalian expression vector was obtained from Florian Krammer (Icahn School of Medicine at Mount Sinai, New York)^25^. This construct was used to transiently transfect 293T cells cultured in Opti-MEM (ThermoFisher Scientific) in 2L roller bottles using Polyethylenimine (PEI) linear (Polysciences, Inc, USA). Supernatant was harvested on day 4 after transfection, dialyzed into PBS overnight, and loaded onto a Ni-NTA agarose (Qiagen) column by gravity flow. The column was washed with PBS containing 10mM imidazole, eluted using 250 mM imidazole in PBS, then buffer exchanged into PBS using a PD10 column (GE Healthcare).

### SARS-CoV-2 spike glycoprotein expression and purification

Expression plasmid encoding SARS-CoV-2 S glycoprotein ^9^ was transiently transfected into Human Embryonic Kidney (HEK) 293F cells. Cells were maintained at a density of 0.2-3 × 10^6^ cells per ml at 37°C, 8% CO2 and 125 rpm shaking in FreeStyle 293F media (Fisher Scientific). Prior to transfection two solutions containing 25 ml Opti-MEM (Fisher Scientific) medium were prepared. Plasmid DNA was added to one to give a final concentration after transfection of 310 μg/L. Polyethylenimine (PEI) max reagent (1 mg/mL, pH 7) was added to the second solution to give a ratio of 3:1 PEI max: plasmid DNA. The two solutions were combined and incubated for 30 minutes at room temperature. Cells were transfected at a density of 1×10^6^ cells per ml and incubated for 7 days at 37°C with 8% CO_2_ and 125 rpm shaking.

After harvesting, the cells were spun down at 4000 rpm for 30 minutes and the supernatant applied to a 500 mL Stericup-HV sterile vacuum filtration system (Merck) with a pore size of 0.22 µm. The supernatant containing SARS-CoV-2 S protein was purified using 5 mL HisTrap FF column connected to an Akta Pure system (GE Healthcare). Prior to loading the sample, the column was washed with 10 column volumes of washing buffer (50 mM Na_2_PO_4_, 300 mM NaCl) at pH 7. The sample was loaded onto the column at a speed of 2 mL/min. The column was washed with washing buffer (10 column volumes) containing 50mM imidazole and eluted in 3 column volumes of elution buffer (300 mM imidazole in washing buffer). The elution was concentrated by a Vivaspin column (100 kDa cut-off) to a volume of 1 mL and buffer exchanged to phosphate buffered saline (PBS).

The Superdex 200 16 600 column was washed with PBS at a rate of 1 mL/min. After 2 hours, 1 mL of the nickel affinity purified material was injected into the column. Fractions separated by SEC were pooled according to their corresponding peaks on the Size Exclusion chromatograms. The target fraction was concentrated in 100 kDa vivaspin (GE healthcare) tubes to ∼1 mL.

### ACE2 expression and purification

To determine the functionality of the purified SARS-CoV-2 S glycoprotein, SPR was performed using truncated soluble angiotensin converting enzyme2 (hACE2). This construct is identical to full length ACE2 except is truncated at position 626. This protein was expressed and purified identically as for the SARS-CoV2 glycoprotein, with the exception of a smaller Vivaspin cutoff being used for buffer exchanging. Following purification, the His-Tag was removed from ACE2 using HRV3C protease cleavage (Thermo Fisher). Digestion was performed at a ratio of 1:20 HRV3C protease: ACE2 in 1x HRV3C reaction buffer (Thermo Fisher) and incubated at 4°C overnight. To remove the HRV3C and uncleaved ACE2 nickel affinity chromatography was performed, except the flow through was collected rather than the elution.

### Surface plasmon resonance (SPR)

After removing metallic contaminants via a pulse of EDTA (350 mM) for 1 min at a flow rate of 30μL/min, the chip was loaded with Ni^2+^ by injecting NiCl_2_ for 1 min at a flow rate of 10 μL/min. SARS-CoV2 S protein (50 μg/mL) was injected at 10μL/min for 3 min. Control channels received neither trimer nor NiCl_2_. Control cycles were performed by flowing the analyte over Ni^2+^-loaded NTA in the absence of trimer; there were no indications of non-specific binding. The analyte was injected into the trimer sample and control channels at a flow rate of 50 μL/min. Serial dilutions ranging from 200 nM to 3.125 nM were performed in triplicate along with HBS P+ buffer only as a control. Association was recorded for 300 s and dissociation for 600 s. After each cycle of interaction, the NTA-chip surface was regenerated with a pulse of EDTA (350 mM) for 1 min at a flow rate of 30 μL/min. A high flow rate of analyte solution (50 μL/min) was used to minimize mass-transport limitation. The resulting data were fit to a 1:1 binding model using Biacore Evaluation Software (GE Healthcare) and these fitted curves were used to calculate K_D_.

### ELISA methodology

96-well high-binding plates (Corning, USA) were coated overnight at 4 °C with antigens at the stated dilutions in sterile PBS. Plates were blocked with 2% BSA (Sigma, UK) prepared in PBS-0.1% Tween 20 for 1 h at room temperature (RT). Pre-diluted serum or saliva samples were added (100 µL per dilution) and serially diluted and plates incubated for 1 h at RT. After washing with PBS-0.1% Tween, 100 µL of HRP-conjugated or unconjugated mouse anti-human immunoglobulins were added and incubated for 1 h at RT. Anti-human immunoglobulin antibodies (anti-IgG, clone R-10 1:8000; anti-IgA clones, 2D7 1:2000 and MG4.156 1:4000; anti-IgM clone AF6 1:2000; anti-IgG1 clone MG6.41, 1:3000; anti-IgG2 clone MG18.02 1: 3000; anti-IgG3 clone MG5.161 1:1000; anti-IgG4 clone RJ4 1:1000, are all clones generated in the University of Birmingham and available from Abingdon Health, UK). In some experiments, HRP-labelled goat anti-mouse immunoglobulins (Southern Biotech, USA) were added and incubated for 1 h at RT. After washing, plates were developed for 5-10 min with 100 µL of TMB core (Biorad, UK) and then stopped with 50 µL of 0.2M H_2_SO_4_. OD was recorded at 450 nm using the Dynex DSX automated liquid handler (Dynex Technologies, USA). Signal:noise ratio (S:N ratio) was calculated by dividing the average OD from the positive samples (signal) over the average OD from the pre2019 negative controls (noise).

### Statistical analysis

Pairwise Pearson’s correlation coefficient was used to calculate the correlation of matched serum and saliva data. The data was classed as positive and negative based on the cut-off established from the concentrations of pre2019 samples assuming that the values of the mean + 3 standard deviations were biologically plausible to be negative for anti-SARS-CoV-2 antibodies (Cutt-off values 0.349 in saliva and 0.629 in serum). The agreement between the classification of saliva and serum samples was assessed using kappa statistics with STATA 16.1 (StataCorp LLC, USA).

## Data Availability

Data is available upon reasonable request from the corresponding author.

## Authors contributions

SEF, SEJ, MPT designed and performed experiments, interpreted results and wrote the manuscript. AMS interpreted results and wrote the manuscript. JDA, YW and MLN designed and performed experiments. AC performed experiments. CRW, MS and MG provided essential reagents. JLH, EMJ and GLM performed experiments. BT interpreted results, DCW interpreted results and wrote the manuscript. TVV provided essential clinical material. SH provided essential reagents. SJ and MJP provided essential clinical material. TP provided experimental support. AH provided essential clinical material. MKO interpreted results. BJE designed experiments and provided essential reagents. MTD designed experiments, interpreted results and wrote the manuscript. MC, AFC and AGR designed experiments, interpreted results, wrote the manuscript and supervised the project. All authors commented on drafts of the manuscript and approved the final version.

## Acknowledgments

We would like to thank the University of Birmingham Clinical Immunology Service for their invaluable support in sample collection and processing. We would also like to thank the National Institute for Health Research (NIHR)/Wellcome Trust Birmingham Clinical Research Facility staff for their vital support in study participant recruitment and sample collection. We also gratefully acknowledge the University of Birmingham Protein Expression Facility for use of mammalian expression equipment. AFC is grateful for funding from The Medical Research Council, The Institute for Global Innovation and The University of Birmingham. This study was supported by the UK National Institute for Health Research, Birmingham Biomedical Research Centres Funding scheme. The work in Prof. Max Crispin’s laboratory was funded by the International AIDS Vaccine Initiative, Bill and Melinda Gates Foundation through the Collaboration for AIDS Vaccine Discovery (OPP1196345/INV-008813, OPP1084519 and OPP1115782), the Scripps Consortium for HIV Vaccine Development (CHAVD) (NIH: National Institute for Allergy and Infectious Diseases AI144462), and the University of Southampton Coronavirus Response Fund. MJP is funded by the Welsh Clinical Academic Training (WCAT) programme and is a participant in the NIH Graduate Partnership Program. The CoCo study was funded internally by the University of Birmingham and University Hospitals Birmingham NHS Foundation Trust and carried out at the National Institute for Health Research (NIHR)/Wellcome Trust Birmingham Clinical Research Facility. The views expressed are those of the authors(s) and not necessarily those of the NHS, the NIHR or the Department of Health.

